# Risk assessment and prediction of severe or critical illness of COVID-19 in the elderly

**DOI:** 10.1101/2020.05.11.20094383

**Authors:** Xiao-Yu Zhang, Lin Zhang, Yang Zhao, Wei-Xia Li, Hai-Bing Wu, Yun Ling, Zhi-Ping Qian, Yin-Peng Jin, Qing-Chun Fu, Xin-Yan Li, Yi Zhang, Yu-Xian Huang, Liang Chen

## Abstract

**Background:** This study aims to investigate the clinical characteristics and risk prediction of severe or critical events of COVID-19 in the elderly patients in China.

**Methods:** The clinical data of COVID-19 in the elderly patients admitted to the Shanghai Public Health Clinical Center during the period of January 20, 2020 to March 16, 2020 were collected. A retrospective cohort study design was conducted to screen out independent factors through Cox univariable regression analysis and multivariable regression analysis, and the efficacy of risk prediction of severe or critical illness was examined through the receiver operating characteristic (ROC) curve.

**Results:** A total of 110 elderly patients with COVID-19 were enrolled. 52 (47.3%) were males and 21 (19.1%) had severe or critical illness. Multivariable regression analysis showed that CD4 cells and D-dimer were independent risk factors. D-dimer, CD4 cells, and D-dimer/CD cells ratios with cut off values of 0.65 (mg/L), 268 (cell/ul) and 431 were in the prediction of severe or critical illness of the elderly COVID-19. The AUC value of D-dimer, CD4 cells, CD4 cells/D-dimer ratio, the tandem group and the parallel group were 0.703, 0.804, 0.794, 0.812 and 0.694, respectively.

**Conclusions:** D-dimer, CD4 cells and their combination have risk assessment value in predicting severe or critical illness of COVID-19 in the elderly.

## 1. Introduction

The disease burdens of coronavirus disease 2019 (COVID-19), caused by severe acute respiratory syndrome coronavirus-2 (SARS-CoV-2), have been continuously increasing [1]. To date, there have been over 3.5 million confirmed patients and over 0.24 million deaths globally according to COVID-19 global data from Johns Hopkins University (JHU). Elderly has been reported as a significant predictor of morbidity and mortality in patients with COVID-19 [2]. The mortality of elderly patients with COVID-19 is higher than that of young and middle-aged patients and elderly patients with COVID-19 are more likely to progress to severe disease [2]. A rennet meta-analysis found that the case-fatality rate (CFR), proportion of severe cases, acute respiratory distress syndrome (ARDS) were significantly higher in studies in which the proportion of elderly patients was larger [3].A single center retrospective study found that elder patients with comorbidities and ARDS were at increased risk of death [4]. A multivariate Cox regression analysis results showed age and severe cases were identified as independent prognostic factors for virus clearance. Another study also confirmed that high proportion of severe to critical cases and high fatality rate were observed in the elderly COVID-19 patients [5]. Furthermore, a study showed that older patients with comorbidities and ARDS are at increased risk of death [4]. Given a large aging population worldwide, it is important to understand characteristics and susceptibility of and impact on elderly patients with COVID-19. Up to now, there has been few reports on risk assessment of severe or critical illness of COVID-19 in the elderly [2, 5, 6]. This study aims to provide a comprehensive understanding clinical characteristics of elderly patients with COVID-19 and determinants of prognosis.

## 2. Materials and Methods

### 2.1 Patients and definitions

Patients with COVID-19 confirmed diagnosis admitted to the Shanghai Public Health Clinical Center from January 20, 2020 to March 16, 2020 were included. Confirmed diagnosis met following criteria: the patients had epidemiological contact history or clinical manifestations, and positive nucleic acid of COVID-19 was confirmed with real-time fluorescence polymerase chain reaction (RT-PCR). Severe events of COVID-19 patients was defined as one of the following conditions: shortness of breath with respiratory rate (RR) ≥30 times/min; the oxygen saturation ≤93% at rest; the arterial partial oxygen pressure (PaO2)/oxygen absorption concentration (FiO2) ≤ 300mmHg. Critical events of COVID-19 patients was defined as one of the following conditions: respiratory failure with the need for mechanical ventilation; shock; patients with other organ-failures should be treated in ICU [7].Elderly patients were defined as aged ≥ 60 years old. Severe or critical illness were defined as patients with COVID-19 developed severe or critical events during hospitalization. The index date was set as the first day of hospitalization. The endpoint was the occurrence of severe or critical illness during hospitalization. The high-risk group was defined as elderly patients with COVID-19 who developed severe or critical events during hospitalization, and the control group was defined as the elderly patients with COVID-19 who did not develop to severe or critical illness during hospitalization.

Informed consents of patients were obtained for diagnosis and treatment, and the study protocol was approved by the Shanghai Public Health Clinical Center Clinical Committee. All the data received Institutional Review Board (IRB) approval by the Ethics Committee. The IRB number was YJ-2020-S015-01.

### 2.2 Study design and indicator*s*

A retrospective cohort study design was conducted to collect the demographic and clinical data from electronic medical record (EMR) including hospital information system (HIS), laboratory information system (LIS) and radiology information system (RIS) of Shanghai Public Health Clinical Center. The demographic and clinical data included age and sex of the patients, the number of pulmonary lobe involved by COVID-19 on the CT scan, history of hypertension, diabetes, cardiovascular disease; laboratory testing results including alanine aminotransferase (ALT), aspartate aminotransferase (AST), creatine kinase (CK), lactate dehydrogenase (LDH), total bilirubin (TBIL), albumin (ALB), prealbumin (PA), total cholesterol (TC), triglyceride (TG), urea (BUN), creatinine (Cr), high-density lipoprotein (HDL-C), low-density lipoprotein (LDL-C), activated partial thrombin time (APTT), fibrinogen (Fg) and thrombin time (PT), coagulation time (TT), D-dimer (D-D), leukocyte (WBC), leukocyte(Neu), lymphocyte(Lym), hemoglobin (Hgb), blood platelet (PLT), and CD4+cells(CD4). All the above baseline information was collected in the 24 hours after admission.

### 2.3 statistical analyses

Person chi-square test was used for counting data analysis. The K-S test was used for the normality of continuous variables. Data with normal distribution were described as mean ± standard deviation. The median and quartile was used for the data without normal distribution. The difference between case and control groups was tested by Manny-Whitney U.

The possible risk factors of severe or critical illness were investigated with Cox proportional hazards (PH) regression models for univariate and multivariate analyses to estimate hazard ratios (HRs) and 95% confidence intervals (CIs). PH assumption was verified using Schoenfeld residuals.

For the prediction indicators, the cut-off value was calculated according to of maximum Youden Index. Youden Index (YDI) is the main summary statistic of the receiver operating characteristic (ROC) curve used in the interpretation and evaluation of an indicator, which defines the maximum potential effectiveness of a diagnostic test. An optimum cut-off point can be determined by calculating Youden’s index (J can be formally defined as YDI = maxc {Se (c) + Sp (c) − 1}) for each possible cut-off point. The cut-off point that achieves this maximum Youden’s Index is referred to as the optimal cut-point (c*) because it is the cut-off point that optimizes the diagnostic test’s differentiating ability when equal weight is given to sensitivity and specificity. The prediction efficiency of prediction indicators was evaluated by ROC curve. The area under curve (AUC) of ROC were compared using Delong test. The P value of two-sided less than 0.05 was considered as statistically significant. SPSS software version 16.0 (SPSS Inc. Chicago, IL, USA,) was used for statistical analysis of the data.

## 3. Results

### 3.1 Baseline profile of COVID-19 in elderly adults

A total of 110 elderly patients of confirmed COVID-19 were included in this study, in which 52 were males (47.3%). 21 patients (19.1%) were included in the high-risk group and 89 patients (80.9%) in the control group. The proportion of males, aspartate aminotransferase, creatine kinase, lactate dehydrogenase, total bilirubin, creatinine, activated partial thrombin time, thrombin time, creatinine, D-dimer, neutrophil count, neutrophil percentage and hemoglobin in the high-risk group were higher than those in the control group (P<0.05). The level of albumin, pre-albumin, lymphocytes and CD4 cells in the high-risk group were lower than those in the control group (P<0.05). T (P<0.05). The detailed information was shown in **Table 1**.

**Table 1.** Baseline data of the elderly patient with COVID-19

| Indicators | Control group | Risk group | $\chi^2/Z/t$ | P |
| --- | --- | --- | --- | --- |
| Number of cases | 89 | 21 |  |  |
| Age (years) | 66.00±6.47 | 71.19±7.83 | 1.933 | 0.053 |
| Sex, male (%) | 35(39.33) | 17(80.95) | 11.812 | 0.001 |
| The onset time (day) | 6.31±3.77 | 6.52±5.56 | 0.207 | 0.836 |
| CT pulmonary infiltration number | 5.00(2.00-5.00) | 5.00(4.00-5.00) | 1.639 | 0.101 |
| Hypertension (n, %) | 32(35.96) | 8(38.1) | 0.034 | 0.854 |
| Diabetes mellitus (n,%) | 17(19.1) | 3(14.29) | 0.040 | 0.841 |
| Cardiovascular disease (n, %) | 13(14.61) | 6(28.57) | 1.445 | 0.229 |
| ALT(U/L) | 20.00±11.2 | 26±10.71 | 1.485 | 0.138 |
| AST(U/L) | 24.00±11.24 | 35.76±13.06 | 2.707 | 0.007 |
| CK(U/L) | 128.8±152.49 | 235.71±159.57 | 2.865 | 0.005 |
| LDH(U/L) | 252.13±74.5 | 384.52±120.11 | 4.836 | 0.000 |
| TBIL(umol/L) | 8.30±4.35 | 12.32±6.22 | 2.529 | 0.011 |
| ALB(g/L) | 39.2±3.21 | 35.15±4.44 | 4.811 | <0.001 |
| PA(mg/L) | 137.62±55.39 | 82.53±36.12 | 4.337 | <0.001 |
| TC(mmol/L) | 4.13±0.87 | 4.13±1.23 | 0.005 | 0.996 |
| TG(mmol/L) | 1.15±0.48 | 1.01±0.32 | 0.259 | 0.796 |
| BUN(mmol/L) | 5.44±1.96 | 8.52±8.68 | 1.613 | 0.122 |
| Cr(umol/L) | 67.42±24.7 | 72.33±28.6 | 2.358 | 0.018 |
| HDL-C(mmol/L) | 1.17±0.3 | 1.03±0.22 | 1.984 | 0.050 |
| LDL-C(mmol/L) | 2.82±0.88 | 2.84±1.19 | 0.109 | 0.913 |
| APTT(sec) | 39.20±8.91 | 43.55±7.95 | 2.095 | 0.036 |
| Fg(g/L) | 4.66±1.19 | 5.15±1.54 | 1.574 | 0.119 |
| PT(sec) | 13.30(13.00-13.70) | 14.06±1.35 | 2.072 | 0.038 |
| TT(sec) | 16.78±1.59 | 17.24±2.01 | 1.134 | 0.259 |
| D-D(mg/L ) | 0.50(0.39-0.93) | 0.93(0.62-1.87) | 3.225 | 0.001 |
| WBC(10<sup>9</sup>/L) | 4.91±1.65 | 6.83±4.72 | 1.839 | 0.080 |
| Neu(10<sup>9</sup>/L) | 3.36±1.49 | 5.62±4.64 | 2.207 | 0.039 |
| Neu(%) | 66.87±10.03 | 78.11±11.6 | 4.481 | 0.000 |
| Lym(10<sup>9</sup>/L) | 1.09±0.4 | 0.84±0.45 | 2.486 | 0.014 |
| Hgb(g/L) | 129.31±12.23 | 138.9±15.8 | 3.049 | 0.003 |
| PLT(10<sup>9</sup>/L) | 172.22±63.83 | 171.1±69.87 | 0.072 | 0.943 |
| CD4(cell/ul) | 483.91±232.93 | 210.05±160.33 | 5.101 | <0.001 |

### 3.2 Risk factors of COVID-19 severe or critical illness events in elderly adults

From the univariate Cox regression analysis, the HR of age, sex, CK, LDH, TBIL, ALB, PA, BUN, Cr, D-dimer, Neu count, Neu percentage, Lym, Hgb and CD4 were 1.105(1.037-1.176), 5.425(1.824-16.138), 1.002(1.000-1.003), 1.008(1.005-1.011), 1.090(1.027-1.156), 0.817(0.751-0.889), 0.973(0.959-0.986), 1.091(1.039-1.145), 1.009(1.004-1.015), 1.217(1.108-1.336), 1.206(1.090-1.335), 1.229(1.116-1.352), 1.099(1.053-1.148), 0.209(0.059-0.735), 1.057(1.020-1.095) and 0.993(0.989-0.996), respectively (P< 0.05). In the multivariate Cox regression analysis suggested that the increase of D-dimer level and the decrease of CD4 cell count were risk factors of occurrence of severe or critical events, with HR of 1.577(1.072-2.320)and 0.993(0.986-0.999), respectively (P<0.05). The detailed Cox regression results were shown in **Table 2**.

**Table 2.** Analysis of risk events indicators in the elderly with COVID-19

| Indicators | Univariate analysis |  | Multivariate analysis |  |
| --- | --- | --- | --- | --- |
|  | HR(95% CI) | P | HR(95% CI) | P |
| Age (years) | 1.105(1.037-1.176) | 0.002* | 1.051(0.955-1.156) | 0.309 |
| Gender, male (%) | 5.425(1.824-16.138) | 0.002* | 2.305(0.530-10.018) | 0.265 |
| The onset time (day) | 1.019(0.916-1.133) | 0.728 |  |  |
| CT pulmonary infiltration number | 1.408(0.963-2.058) | 0.078 |  |  |
| Hypertension (%) | 1.093(0.453-2.638) | 0.843 |  |  |
| Diabetes mellitus (%) | 0.743(0.219-2.522) | 0.634 |  |  |
| Cardiovascular disease (%) | 2.12(0.822-5.469) | 0.120 |  |  |
| ALT(U/L) | 1.007(0.981-1.033) | 0.609 |  |  |
| AST(U/L) | 1.018(0.998-1.039) | 0.082 |  |  |
| CK(U/L) | 1.002(1.000-1.003) | 0.007* | 1.001(0.998-1.004) | 0.479 |
| LDH(U/L) | 1.008(1.005-1.011) | 0.000* | 1.002(0.996-1.009) | 0.536 |
| TBIL(umol/L) | 1.090(1.027-1.156) | 0.005* | 0.989(0.902-1.085) | 0.814 |
| ALB(g/L) | 0.817(0.751-0.889) | <0.001* | 0.978(0.802-1.193) | 0.827 |
| PA(mg/L) | 0.973(0.959-0.986) | <0.001* | 0.987(0.968-1.007) | 0.209 |
| TC(mmol/L) | 0.989(0.621-1.573) | 0.989 |  |  |
| TG(mmol/L) | 1.357(0.732-2.518) | 0.332 |  |  |
| BUN(mmol/L) | 1.091(1.039-1.145) | <0.001* | 0.999(0.766-1.303) | 0.994 |
| Cr(umol/L) | 1.009(1.004-1.015) | 0.001* | 0.998(0.968-1.029) | 0.909 |
| HDL-C(mmol/L) | 0.189(0.033-1.063) | 0.059 |  |  |
| LDL-C(mmol/L) | 1.011(0.632-1.616) | 0.965 |  |  |
| APTT(sec) | 1.019(0.988-1.050) | 0.230 |  |  |
| Fg(g/L) | 1.244(0.931-1.663) | 0.140 |  |  |
| PT(sec) | 1.159(0.960-1.398) | 0.125 |  |  |
| TT(sec) | 1.120(0.912-1.376) | 0.280 |  |  |
| D-D(mg/L) | 1.217(1.108-1.336) | <0.001* | 1.577(1.072-2.320) | 0.021<sup>#</sup> |
| WBC($10^9/L$) | 1.206(1.090-1.335) | <0.001* | 6.406(0.700-58.603) | 0.100 |
| Neu($10^9/L$) | 1.229(1.116-1.352) | <0.001* | 0.095(0.007-1.232) | 0.072 |
| Neu(%) | 1.099(1.053-1.148) | <0.001* | 1.077(0.950-1.222) | 0.247 |
| LYM($10^9/L$) | 0.209(0.059-0.735) | 0.015* | 2.309(0.349-15.273) | 0.385 |
| Hgb(g/L) | 1.057(1.020-1.095) | 0.002* | 1.015(0.967-1.065) | 0.556 |
| PLT($10^9/L$) | 1.000(0.993-1.007) | 0.999 | | |
| CD4(cell/ul) | 0.993(0.989-0.996) | <0.001* | 0.993(0.986-0.999) | 0.023<sup>#</sup> |

\* In the univariate analysis, $P < 0.05$ was considered as statistically significant, therefore the variates should be considered to include in the multivariate analysis.
<sup>#</sup> In the multivariate analysis, $P < 0.05$ was considered as statistically significant.

### 3.3 Prediction efficacy of D-dimer, CD4 cells and their ratios in the occurrence of severe or critical illness in elderly adults

According to the Jordan index maximization principle, the cut-off values of D-dimer, CD4 cells and CD4 cells/D-dimer ratios were 0.65 (mg/L), 268 (cell/ul) and 431 in the prediction of severe or critical events in elderly COVID-19. In the tandem group, the D-dimer was higher than 0.65mg/L and the CD4 cells were less than 268 cell/ul. In the parallel group, the D-dimer was higher than 0.65mg/L, or the CD4 cells were less than 268 cell/ul. The AUC values of D-dimer, CD4 cell, CD4 cells/D-dimer ratio, the tandem group and the parallel group were 0.703, 0.804,0.794, 0.812, and 0.694, respectively (P<0.05). Among them, the AUC value of D-dimer was lower than that of CD4 cells; In the tandem group, the AUC value of CD4 cells/D-dimer ratio was greater than that of the parallel group, and the AUC value of CD4 cells, CD4 cells/D-dimer ratio and the tandem group was greater than that of the parallel group (P<0.05). **Table 3** and **Figure 1** showed the detailed information.

**Table 3.**
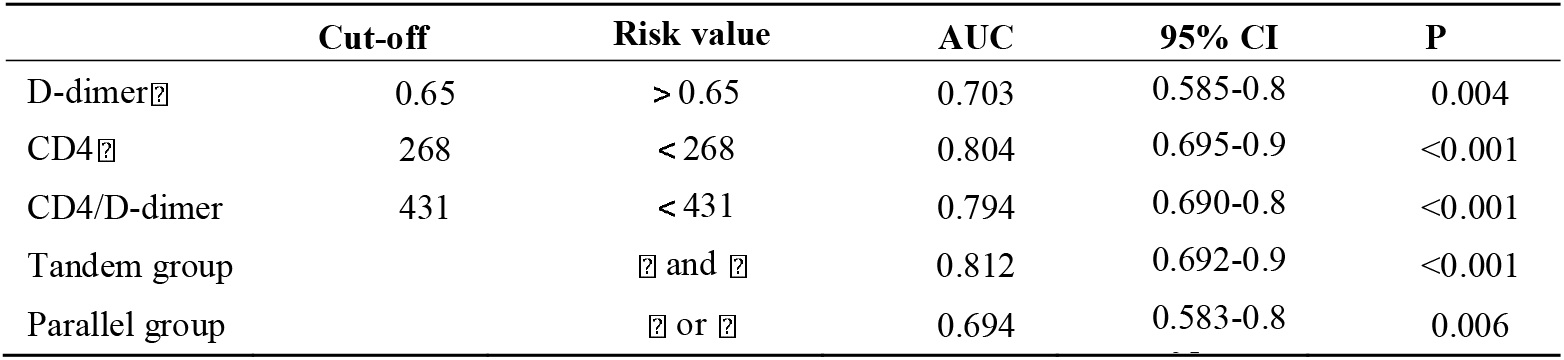
Risk prediction of D-dimer, CD4 cells and their combination in the elderly COVID-19

|  | Cut-off | Risk value | AUC | 95% CI | P |
| --- | --- | --- | --- | --- | --- |
| D-dimer | 0.65 | $> 0.65$ | 0.703 | 0.585-0.8 | 0.004 |
| CD4 | 268 | $< 268$ | 0.804 | 0.695-0.9 | $<0.001$ |
| CD4/D-dimer | 431 | $< 431$ | 0.794 | 0.690-0.8 | $<0.001$ |
| Tandem group | | and | 0.812 | 0.692-0.9 | $<0.001$ |
| Parallel group |  | or | 0.694 | 0.583-0.8 | 0.006 |

**Figure 1.**
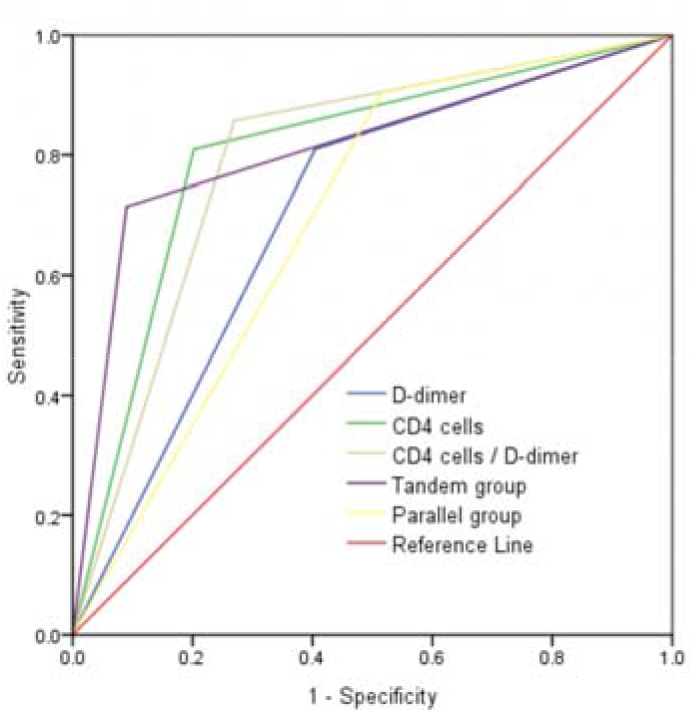
AUC curve of D-dimer, CD4 cells and their combination in the risk prediction of severe or critical illness in the elderly with COVID-19

To evaluate the risk prediction efficacy of COVID-19 in the elderly patients, the sensitivity, specificity, positive predictive value, negative predictive value and Youden Index of D-dimer, CD4 cells, CD4 cells/D-dimer ratio, tandem group and parallel group were showed in **Table 4** with detailed information.

**Table 4.** Evaluation of D-dimer, CD4 cells, and their combination in the risk prediction of severe or critical illness in the elderly with COVID-19

|  | Sen | Spe | PPV | NPV | YDI |
| --- | --- | --- | --- | --- | --- |
| D-D<sup>Ⓢ</sup> | 0.810 | 0.596 | 0.321 | 0.930 | 0.405 |
| CD4<sup>Ⓢ</sup> | 0.810 | 0.798 | 0.486 | 0.947 | 0.607 |
| CD4/D-D | 0.857 | 0.730 | 0.429 | 0.956 | 0.587 |
| Tandem group | 0.714 | 0.910 | 0.652 | 0.931 | 0.624 |
| Parallel group | 0.905 | 0.483 | 0.292 | 0.956 | 0.388 |

Sen:Sensitivity; Spe:Specificity; PPV:Positive predictive value; NPV: Negative predictive value; YDI: Youden Index.

## 4. Discussion

The novel coronavirus is a virus of the family coronavirus, which is derived from bats according to gene sequencing. It belongs to a new independent evolution branch of coronavirus, and has high homology with coronavirus related to severe acute respiratory syndrome (SARS) and coronavirus related to Middle East Respiratory Syndrome (MERS) [8-16]. The virus is mainly spread by droplets and contact, and there is the possibility of aerosol transmission. Incubation patients and asymptomatic virus carriers are also infectious [17].As SARS virus through S protein and angiotensin conversion enzymellreceptor, COVID-19 virus cross-species transmission or human to human transmission [18-20]. Compared with the SARS virus, COVID-19 virus is less virulent but more infectious [19, 20].

Studies have confirmed that aging is an independent contributor to COVID-19 death. Therefore, it is of great value to analyze the risk prediction indicators. This study is mainly to analyze the risk factors and practical applications of severe or critical events in the elderly patients. In this paper, the regression analysis was used to collect COVID-19 keywords from recent literature retrieval, and representative indicators were selected for statistical analysis based on clinical experience and practical value. To avoid the time difference between baseline data and severe or critical events at admission, Cox regression was used for progressive analysis to determine whether the study parameters were independent influencing factors. The ROC curve was used to deduce the warning value of risk factors.

The lung accumulation range of COVID-19 in the early stage was generally wide in the elderly, and the median of both the risk group and the control group was 5 lobes, showing no statistically significant difference between the two groups. Age and sex were the risk factors for COVID-19 severe or critical events in the elderly. The complications of hypertension, diabetes and cardiovascular disease did not significantly aggravate with the severe or critical events. These imported cases may have milder comorbidities; because the cases with severe comorbidities may be less likely to be infected due to the limited range of daily communication.

Subsequently, the sample size could be increased to stratify the severity and control of complications and further verify the risk ratio. However, the number of complications in the elderly is more than that in the non-elderly, and the complications of basic diseases in the elderly is significantly correlated with the occurrence of end-stage event [2, 6].Alanine aminotransferase and aspartate aminotransferase had no effect on severe or critical events in elderly COVID-19, which was consistent with earlier reports [21].Early indicators such as creatine kinase, lactate dehydrogenase, bilirubin, urea, creatinine, albumin, prealbumin, white blood cells, neutrophils and hemoglobin were related factors influencing the severe or critical events of COVID-19 in the elderly, rather than independent factors. It could be related to the basic metabolism, characteristics of infection, body immunity, combination of basic diseases and starting point set by the research in the elderly patients.

In this study, CD4 cells were independent factors affecting severe or critical events of COVID-19 in the elderly. Previous studies of SARS have also confirmed a decrease in the absolute count of CD4 cells, with characteristic changes throughout the pathogenesis, which is consistent with the characteristics of coronavirus family infection. Recent studies have reported that novel coronavirus infection can induce CD4 cell apoptosis [22], Compared with the survival group, the absolute count level of CD4 cells in the death group of viral pneumonia was significantly reduced, suggesting that a large number of T cells were activated and depleted during the antiviral process. All of the above supports the study results of CD4 cells; and CD4 cells < 268 cell/ul have a significant predictive effect on the occurrence of severe or critical COVID-19 events in the elderly, with high sensitivity(81.0%) and negative predictive value (94.7%).

As for the lymphocyte count, there were significant differences in the univariate analysis, but no significant differences in the multivariate analysis. Lymphocyte count was preliminarily determined to be a risk factor associated with COVID-19 severe or critical events in the elderly, rather than an independent factor. Recently, novel coronavirus pneumonia has been reported to have a significant decrease in lymphocytes in the whole population [23, 24]; novel coronavirus may act on lymphocytes via ACE2 receptor, causing lymphocytopenia [25]or induce lymphocytopenia by TNF, interleukin-6, and other pro-inflammatory cytokines [26]; which is also characteristic of coronavirus infection of immune cells. In this study, lymphocytes were indeed low in the elderly, with a lower value of 0.84±0.45 (10^^^9/L) in the risk group, but a normal range of 0.8-3.5 (10^^^9/L). However, the early count of lymphocytes was not an independent factor affecting severe or critical events, which may be related to the characteristics of the elderly. But, studies have suggested that the dynamic expression of lymphocyte percentage can predict the severity of COVID-19 [27]. Later cases could be added, and lymphocyte counts at different stages were used to evaluate the severity of COVID-19 and verify whether there was a statistical difference.

D-dimer was also an independent factor affecting severe or critical events of COVID19 in the elderly. In terms of this indicator, recent studies have reported that the significant increase of D-dimer is associated with the poor prognosis of severe new coronary pneumonia [28, 29]. The death cases of COVID-19 had significant increase of plasma D-dimer [29]. Moreover, in adults with community-active pneumonia, increased plasma D-dimer was associated with increased inflammatory response, admission to intensive care and 30-day mortality, and was superior to C-reactive protein and procalcitonin in predicting admission to intensive care and 30-day mortality [30]. All of the above reflected the value of D-dimer and supported that the increase of D-dimer was an independent factor affecting the severe or critical events of COVID 19 in the elderly. Similarly, the severe or critical events of D-dimer>0.65 mg/L in elderly COVID-19 were also evaluated by ROC curve with high predictive efficacy, with high sensitivity(81.0%) and negative predictive value(93.0%), respectively.

In order to consider the effects of novel coronavirus on immunity and coagulation in elderly patients, we can combined use of D-dimer level and CD4 cell count, to evaluate the occurrence of severe or critical events. The combined use of the both had high inferential efficiency on the occurrence of risk events in the elderly COVID-19, there were respective significant contributions on the evaluation index, with high sensitivity (90.5%), specificity (91.0%), positive predictive value (65.2%) or negative predictive value (95.6%).

Based on the results of this study and the close correlation between autoimmune disorders and coagulation abnormalities and thrombosis events in patients[31], we propose a hypothesis: CD4 cells and D-dimer may trigger in the progression of the elderly COVID-19; novel coronavirus may act on CD4 cells through ACE2 receptor, and affect CD4 cells count reduction; the level of D-dimer can be increased by immune imbalance and inflammatory response in the elderly. In the early stage of the disease, when the number of CD4 cells decreases to a certain number and the D-dimer increase to a certain level, the risk event trend of COVID-19 will be continuously affected. Therefore, the immune imbalance, cytokine disorder and multi-organ damage can be continuously induced in the state of low cellular immunity and high coagulation to a certain extent, leading to the severe or critical events of COVID-19.

The highlight of this study was the elderly COVID-19 as the research object, because the elderly population has its own characteristics with risk factors of the severe or critical events [2, 4, 6]. It was found that D-dimer and CD4 cells were independent factors affecting the severe or critical events of COVID-19 in the elderly, and the indicators had important clinical application value. However, the proportion of comorbidities and their prognostic analysis were significantly different from the other studies [5, 6]. which would be related to the fact that the samples of this study were all imported domestic cases and regional prevention and control policies. The advantage of this study lied in the in-depth analysis of all the research data; the independent influencing factors of the elderly COVID-19 risk events in the elderly were identified, and the application value was evaluated; the study population were all imported domestic cases, and the results were stable. One of the limitations is that data on novel coronavirus load were not available.

## 5. Conclusions

COVID-19 risk factors in the elderly have their characteristics. The decrease of CD4 cell count and the increase of D-dimer level in the early stage of the disease are independent influencing factors for the occurrence of severe or critical events. The application of D-dimer, CD4 cells and the combination have important risk prediction efficacy for the severe or critical illness of the elderly with COVID-19.

## Data Availability

This study was supported by the Shanghai Public Health Clinical Center for data access.

## Author Contributions

Conceptualization, Xiao-Yu Zhang, Wei-Xia Li, Hai-Bing Wu, Yun Ling, Zhi-Ping Qian, Yin-Peng Jin, Qing-Chun Fu, Xin-Yan Li, Yi Zhang, Yu-Xian Huang and Liang Chen; Formal analysis, Xiao-Yu Zhang, Yi Zhang and Yu-Xian Huang; Methodology, Lin Zhang, Yang Zhao, Wei-Xia Li, Hai-Bing Wu, Yun Ling, Zhi-Ping Qian, Yin-Peng Jin, Qing-Chun Fu and Xin-Yan Li; Supervision, Liang Chen; Writing - original draft, Xiao-Yu Zhang, Lin Zhang, Wei-Xia Li, Hai-Bing Wu, Yun Ling, Zhi-Ping Qian, Yin-Peng Jin, Qing-Chun Fu, Xin-Yan Li, Yu-Xian Huang and Liang Chen; Writing - review & editing, Lin Zhang, Yang Zhao, Yi Zhang, Yu-Xian Huang and Liang Chen. All authors have read and agreed to the published version of the manuscript.

## Acknowledgments

This study was supported by the Shanghai Public Health Clinical Center for data access.

## Ethical approval

Informed consents of patients were obtained for diagnosis and treatment, and the study protocol was approved by the Shanghai Public Health Clinical Center Clinical Committee. All the data received Institutional Review Board (IRB) approval by the Ethics Committee. The IRB number was *YJ-2020-S015-01*.

